# Human-Centered Design and Evaluation of an LLM-based Decision Support Tool for Nurses in Rural India

**DOI:** 10.64898/2026.11.23.26368760

**Authors:** Priyank Jain, Theodore Bolas, Sarah Nabia, Anup Agarwal, Chetanya Malik, Abhitesh Tripathi, Shaurya Sharma, Priyansh Bansal, Aadarsh Agarwal, Aarjav Jain, Arnav Malhotra, Rubayat Khan, Abhijit Chowdhury, Parthasarathi Mukherjee

## Abstract

Large language models (LLMs) perform well on physician reasoning tasks, sparking interest in using them to guide mid-level health providers where physicians are scarce. However, evidence is lacking for effective LLM interfaces and their performance in real patient encounters. We report the human-centered design and performance of the NurseAI mobile app in rural West Bengal, India. Nurses participated in six design rounds that culminated in an audio-based mobile app that listens to the patient interview in the patient’s native language, suggests patient-specific questions for the nurse, and generates an assessment and plan. Physician graders evaluated the app’s output for fifty real-patient consultations in the final version and found 90% of the plans were appropriate, 96% valid, and 94% safe. Several feature improvements have been identified for future versions of the app from the field use.

## 1. Introduction

More than half the world’s rural population lives in areas with poor access to doctors^1^. In rural India, the physician-to-resident ratio is nearly four times lower, and the life expectancy is three years less than in urban areas^2^. The Government of India launched the Ayushman Bharat initiative to provide universal, free, comprehensive primary care through a network of 150,000 rural health centers called Ayushman Aarogya Mandirs (AAMs)^3^. A mid-level health provider (MLHP) called a Community Health Officer (CHO) leads a team of non-physician healthcare workers at AAMs. CHOs are either nurses or *AYUSH* (Ayurveda, Yoga and Naturopathy, Unani, Siddha, and Homeopathy) practitioners with six months of dedicated training in community and primary health care^4^. A telemedicine portal called e-Sanjeevani supports the CHOs by connecting them to physicians at higher centers. However, publicly available data on e-Sanjeevani utilization and quality of care suggests a disproportionately low utilization of the tool from rural primary care settings^5^. Low levels of provider knowledge and quality of practice are also well described in rural Indian primary care settings^6–8^.

The rapid growth in capability and accessibility of generative artificial intelligence (AI) has led to an explosion of research studying its clinical applications. Large language models (LLMs) have passed medical licensing exams and surpassed physicians in many diagnostic and management reasoning tasks^9–12^. Evidence is accumulating from low- and middle-income countries (LMICs) that LLMs can augment physician clinical reasoning and can match or exceed physicians’ advice to frontline nurses^9,13^. However, most of these studies have used vignette-based benchmarks, and the positive results are yet to be reproduced with real patients. In a recent cluster-randomized trial in Kenya, non-physician clinicians using AI to assist with treatment decisions demonstrated no significant differences in short-term patient outcomes despite better-quality treatment plans^14^. Translating strong AI performance to real-world settings is also complicated as a recent meta-analysis found that human–AI combinations often perform worse than humans or AI alone^15^. However, careful design of the human-computer interaction may enhance physician-plus-AI performance^16^.

Even if AI-enabled clinical decision support systems (AI-CDSS) are effective, they are useless without user adoption^17,18^. Usability, workflow misalignment, and environmental barriers are the major challenges preventing the effective use of AI-CDSS in clinical settings^19^. Human-centered design (HCD) addresses these sociotechnical challenges by prioritizing user experience and local organizational complexity to improve adoption and sustainability^20,21^. Ninety-eight percent of India’s villages are within mobile network coverage, making mobile phones an ideal platform for frontline healthcare workers to access AI-CDSS^22^.

Nurses and midwives constitute 39% of the country’s health workforce and are often tasked with delivering primary healthcare services^23^. Given the shortage of doctors in rural India and AI’s growing clinical capabilities, equipping MLHPs with AI may bridge healthcare gaps in areas with physician understaffing. Thus, we conducted a randomized crossover trial (NCT07432893) to assess the effectiveness of AI-enabled nurse-led primary care in rural India. This paper does not report the results of this study, which are forthcoming, but rather aims to (1) describe the design process of developing an LLM-based CDSS for nurses (a type of MLHP) to conduct clinical consultations in a rural primary care setting, and (2) report the evaluation of the NurseAI app (“the app”) in real patient encounters.

## 2. Results

### 2.1 Human-Computer interaction design

Our search for an optimal user interface for AI started from the simplest method: a commercially available general-purpose chatbot. We then iterated based on feedback from nurses and observations from physician trainers. We tested six main iterations detailed in Table 1. The first four prototypes were tested using clinical role-play and were not used with real patients. The final two iterations were versions of the mobile app, named NurseAI version 1 (v1) and NurseAI version 2 (v2). Version 1 was the minimum viable product deployed on 25 December 2025 for pilot patients and v2 was deployed on 13 January 2026 for study patients.

**Table 1:**
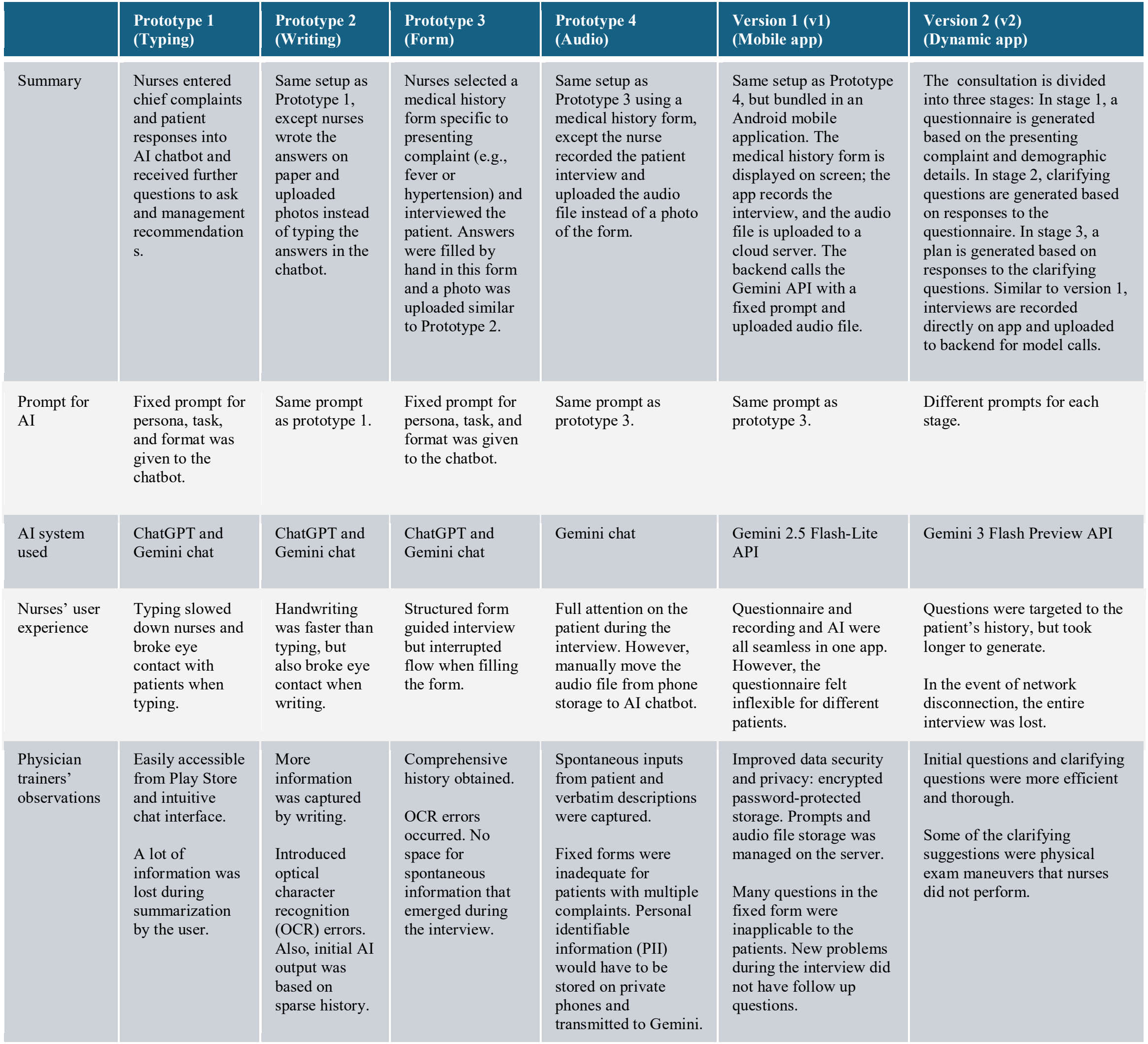
Comparison of Nurse and AI interface iterations.

|  | **Prototype 1<br>(Typing)** | **Prototype 2<br>(Writing)** | **Prototype 3<br>(Form)** | **Prototype 4<br>(Audio)** | **Version 1 (v1)<br>(Mobile app)** | **Version 2 (v2)<br>(Dynamic app)** |
|---|---|---|---|---|---|---|
| Summary | Nurses entered chief complaints and patient responses into AI chatbot and received further questions to ask and management recommendations. | Same setup as Prototype 1, except nurses wrote the answers on paper and uploaded photos instead of typing the answers in the chatbot. | Nurses selected a medical history form specific to presenting complaint (e.g., fever or hypertension) and interviewed the patient. Answers were filled by hand in this form and a photo was uploaded similar to Prototype 2. | Same setup as Prototype 3 using a medical history form, except the nurse recorded the patient interview and uploaded the audio file instead of a photo of the form. | Same setup as Prototype 4, but bundled in an Android mobile application. The medical history form is displayed on screen; the app records the interview, and the audio file is uploaded to a cloud server. The backend calls the Gemini API with a fixed prompt and uploaded audio file. | The consultation is divided into three stages: In stage 1, a questionnaire is generated based on the presenting complaint and demographic details. In stage 2, clarifying questions are generated based on responses to the questionnaire. In stage 3, a plan is generated based on responses to the clarifying questions. Similar to version 1, interviews are recorded directly on app and uploaded to backend for model calls. |
| Prompt for AI | Fixed prompt for persona, task, and format was given to the chatbot. | Same prompt as prototype 1. | Fixed prompt for persona, task, and format was given to the chatbot. | Same prompt as prototype 3. | Same prompt as prototype 3. | Different prompts for each stage. |
| AI system used | ChatGPT and Gemini chat | ChatGPT and Gemini chat | ChatGPT and Gemini chat | Gemini chat | Gemini 2.5 Flash-Lite API | Gemini 3 Flash Preview API |
| Nurses’ user experience | Typing slowed down nurses and broke eye contact with patients when typing. | Handwriting was faster than typing, but also broke eye contact when writing. | Structured form guided interview but interrupted flow when filling the form. | Full attention on the patient during the interview. However, manually move the audio file from phone storage to AI chatbot. | Questionnaire and recording and AI were all seamless in one app. However, the questionnaire felt inflexible for different patients. | Questions were targeted to the patient’s history, but took longer to generate.<br>In the event of network disconnection, the entire interview was lost. |
| Physician trainers’ observations | Easily accessible from Play Store and intuitive chat interface.<br>A lot of information was lost during summarization by the user. | More information was captured by writing.<br>Introduced optical character recognition (OCR) errors. Also, initial AI output was based on sparse history. | Comprehensive history obtained.<br>OCR errors occurred. No space for spontaneous information that emerged during the interview. | Spontaneous inputs from patient and verbatim descriptions were captured.<br>Fixed forms were inadequate for patients with multiple complaints. Personal identifiable information (PII) would have to be stored on private phones and transmitted to Gemini. | Improved data security and privacy: encrypted password-protected storage. Prompts and audio file storage was managed on the server.<br>Many questions in the fixed form were inapplicable to the patients. New problems during the interview did not have follow up questions. | Initial questions and clarifying questions were more efficient and thorough.<br>Some of the clarifying suggestions were physical exam maneuvers that nurses did not perform. |

### 2.2 The app workflow

In both versions, the nurse registers the patient and the app records the interview. In v1, the nurse asks the questions on a fixed medical history form chosen for the presenting complaint (e.g., fever), and the app returns an assessment and plan. In v2, the nurse can instead ask the app to generate a patient-specific questionnaire from the patient’s age, sex, presenting symptoms, and other concerns (e.g., fever and abdominal pain in a 25-year-old woman). Based on the answers to this questionnaire the nurse receives clarifying questions and physical examination to narrow the differential, and in the next step receives th assessment and plan (Figure 1; Table 1). The technical stack is described in Methods section 4.4.

**Figure 1.**
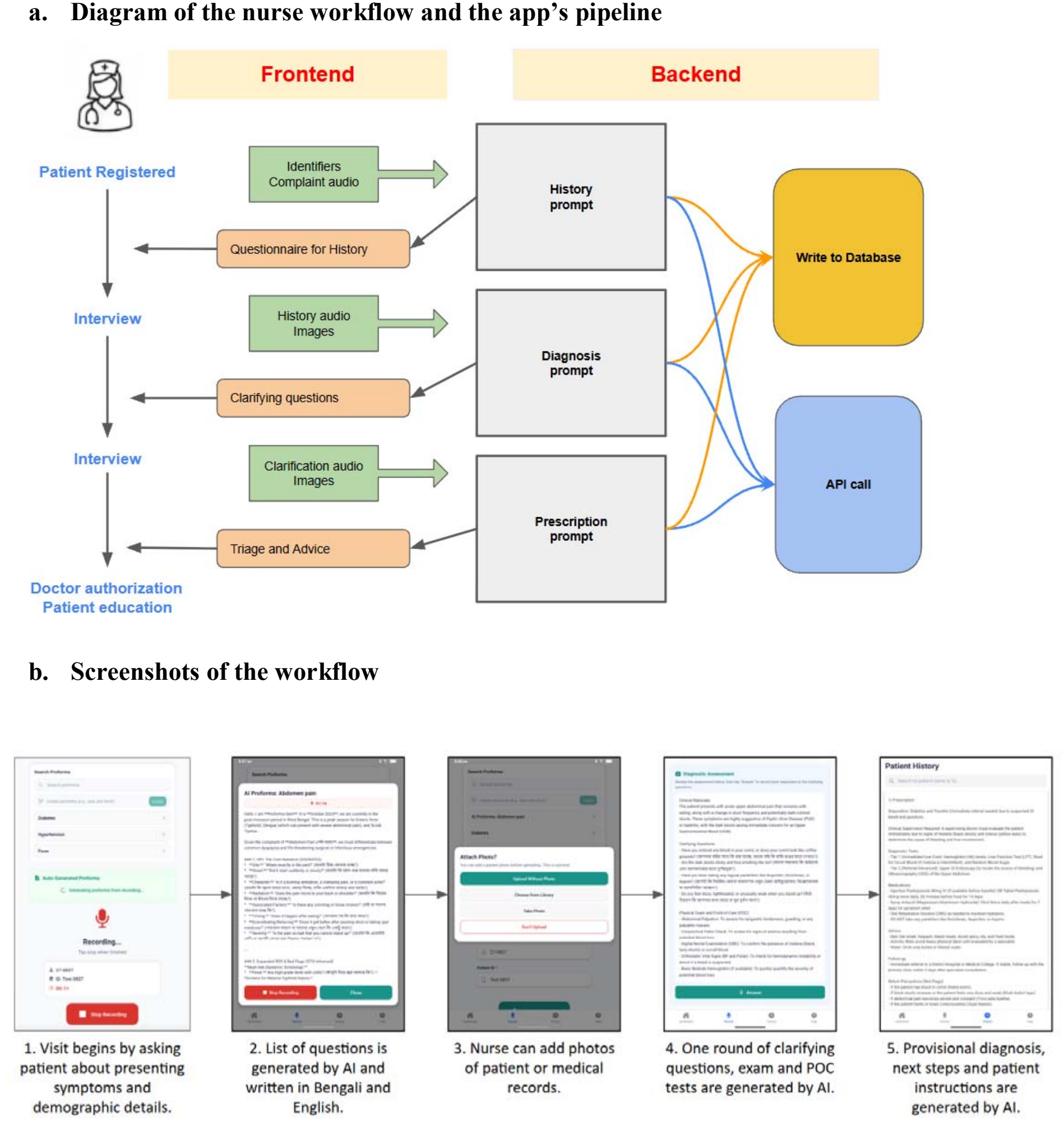
NurseAI app version 2.

### 2.3 Technical and clinical evaluation

Physician graders independently rated the assessment and plan of fifty randomly selected patient consultations from v2. They gave identical ratings on all three criteria in 80% cases (40 of 50). The technical and clinical evaluation results are summarized in Table 2.

**Table 2:** Technical and Clinical evaluation of version 2.

| Technical measures (n = 50)* | Version 2 |
|---|---|
| Interview length, mean (sd; range) | 7m 24s (1m 43s; 3m 35s-12m 31s) |
| Latency per call, mean (sd), seconds* | Questionnaire: 21.6 (3.5)<br>Diagnosis: 12.9 (2.2)<br>Prescription: 12.4 (1.8) |
| Latency per consultation, mean (sd), seconds* | 46.9 (4.4) |
| Tokens per consultation, mean (sd) | 22,230 (2,656) |
| Token cost per consultation, mean (sd), US cents | 3.5 (0.4) |
| Clinical measures (n=50) |  |
| Appropriate, n (%) [95% CI] | 45 (90%) [78.2, 96.7] |
| Valid, n (%) [95% CI] | 48 (96%) [86.3, 99.5] |
| Safe, n (%) [95% CI] | 47 (94%) [83.5, 98.7] |

\*does not include upload times

The assessment and plan was rated appropriate in 90% of the cases. Overinvestigation was the most common error. Examples included suggesting complete blood count (CBC) for upper respiratory illness (URI) or urine microscopy and serum creatinine for acute diarrhea which were considered unnecessary. Some suggestions like Brucella rapid test card or vitamin b12 level were unavailable in rural health center settings.

The assessment and plan was rated valid in 96% of the cases. The two errors were because of implausible diagnoses included in the differentials. One was alcohol neuropathy listed as a cause for acute low back pain that originated after lifting a heavy object. The other was malaria listed as a possible explanation for a two-month duration of fever and weight loss.

The assessment and plan was rated safe in 94% of the cases. All the unsafe plans were because of errors of omission rather than harmful recommendations. These included failure to consider must-not-miss diagnoses such as tuberculosis in a case of two-month duration of fever and weight loss, not suggesting EKG in a young woman with chronic breathlessness and chest pain, and not suggesting antiplatelet or statins for stroke prevention in one case of post-stroke patient presenting with subacute weakness and swelling.

## 3. Discussion

We demonstrate that an iterative, human-centered design process resulted in an audio-based mobile app that guides a nurse through a clinical consultation. With v2, the interview alone averaged nearly seven and a half minutes, longer than the 2-minute average recorded for entire consultations in previous studies of Indian primary care^24^. It cost an average of 3.5 cents and produced an assessment and plan with encouraging validity, safety, and appropriateness scores. To our knowledge, this is among the first AI-enabled CDSSs to support mid-level health providers in completing an entire patient consultation using audio recordings in a native language.

Nurse users preferred a voice interface over typing so we chose a general-purpose LLM (Gemini) over healthcare-specific LLMs because of its multimodal and multilingual capabilities. Physician trainers observed that history-taking was initially limited to 3–4 questions, consistent with studies of public-sector providers in rural India^7,25,26^. This informed training sessions on medical interviewing and using medical history forms to gather richer context for AI guidance.

Version 2 introduced four modifications: two design and two technical. The two design modifications were adding a dynamic, patient-specific questionnaire generated from the patient’s chief complaints and a round of clarifying questions generated from the initial interview. The two technical modifications were updated prompts and a change from Gemini 2.5 Flash-Lite to Gemini 3 Flash Preview. After using Version 1 with real patients, nurses reported that patients typically presented with more than one symptom, and history proformas for a single symptom or disease were not adequate for taking a complete history. This feedback drove the design change from using fixed proformas in the app to a dynamic, patient-specific history-taking feature. A hypothetico-deductive approach to diagnostic reasoning uses follow-up questions to clarify and reorder the differential^27^. The clarifying-question step of v2 was designed to support this, similar to other conversational diagnostic systems^28^.

Grader comments from clinical evaluation have identified areas for improvement. The most common appropriateness error (four of five errors) was a mismatch with the socioeconomic context. Graders found that the app frequently recommended tests such as CBC and chest X-rays that are not easily available in rural Indian settings. These errors occurred despite adding guideline-directed and cost-conscious prompt instructions in v2. This suggests that prompt engineering alone may be insufficient to reliably constrain an LLM to what is appropriate for a rural-Indian setting. Consequently, the app’s next version adds retrieval-augmented generation from the Indian Council of Medical Research (ICMR) Standard Treatment Workflows^29^. All safety errors (three errors) in version 2 involved omissions rather than harmful recommendations, and our findings are consistent with the NOHARM benchmark, in which omissions accounted for more than 80% of severe safety errors across LLMs and clinical AI tools^30^. Omissions may be harder to catch than harmful recommendations even with a human in the loop. We therefore plan to add a safety gate that checks each plan against a deterministic list of must-not-miss diagnoses and red flags.

Feedback from nurses after real-world use of the version 2 during the randomized crossover trial has provided insights into areas for further improvement. First, nurses reported varying preferences for the script used to display text in the app. Second, users reported wanting more control over parts of the interview they do not want to record because of sensitivity and privacy concerns around the patient’s emotional details, which have no bearing on the clinical aspects of the case. Third, using the app in remote, rural settings in India caused slow and dropped uploads due to patchy internet connectivity.

Other AI applications for frontline health workers in LMICs include AIRA by Qure.ai in India^31^, Ayu 2.0 by Intelehealth in India^32^, AIFYA in Kenya^33^, SCIP in Ethiopia^34^, and an EHR-integrated tool from Penda Health in Kenya^35^. AIRA is an LLM-based platform that develops patient summaries from audio conversations and generates protocol-based nudges and program-level intelligence. Ayu 2.0 guides health workers through protocol-based patient history capture and sends AI-generated diagnosis and management plans to the supervising physician^32^. AIFYA and SCIP are an LLM-based clinical decision support system that answer clinician’s questions based on selected knowledge corpus. Another LLM-based solution tested in an LMIC is by Penda Health in Kenya, where an EHR-integrated AI solution provides alerts and recommendations based on patient data entered in the EHR^35^. All these apps rely on the frontline clinician’s ability to acquire the relevant clinical information, summarize it, and formulate the clinical question for guidance. Compared with these other products, the NurseAI app’s differentiating characteristic is its AI-first workflow. AI helps gather patient information and provides it as model context, along with carefully adjusted system prompts that elicit clinical guidance from the LLM. Information gathering is not solely left to the clinician’s skills.

A comparison of the participatory design process of our app to other AI-CDSSs built in LMICs revealed sparse evidence from varied settings but full-cycle examples are rare^36–38^. One report from Cambodia offers a concrete iterative design like our process, although it was for a non-AI CDSS^39^. A key difference in our HCD approach was that we framed the problem upfront through a research question on AI-enabled nurse-led consultations, and the frontline users remained involved throughout the rest of the HCD cycle.

This documentation and testing process is not without drawbacks. First, we did not formally evaluate and qualitatively analyze each iteration, and each iteration bundled multiple changes, so we cannot isolate the impact of individual modifications. Second, patients did not have direct input in the design process, especially regarding the provider-patient interaction. Third, grading an assessment and plan for real patients is challenging because ground truth can only be obtained through case follow-up and confirmatory tests, neither of which is readily available in a low-resource setting. Finally, we could not assess bias in the app’s outputs because we did not routinely collect caste, ethnicity, or religion during consultations.

In conclusion, a human-centered design process with nurses resulted in an audio-based LLM decision support app that may be safe, appropriate, and valid in a rural Indian setting. Because the app does not rely on the user’s ability to formulate questions relevant to a patient’s condition, it can be used by other MLHPs in similar settings. Whether the app improves patient outcomes remains an important open question that requires further research with health outcome endpoints.

## 4. Methods

### 4.1 Ethics approval

This study followed the principles outlined in the Declaration of Helsinki. The Institutional Ethics Committee of the Indian Institute of Liver and Digestive Sciences (IILDS) approved the study prior to participant enrollment (approval reference IILDS/2025-R70). We obtained written informed consent from all participants before any study procedures. This paper was written in accordance with the DECIDE-AI reporting guidelines, and a completed checklist is available in the Supplementary Information. User learning curves were not formally measured in this study.

### 4.2 Context

#### Study setting

This study was conducted in Birbhum and Purulia districts in West Bengal, India. These areas are primarily Bengali speaking, with agriculture as the largest source of employment and literacy rates of 65-70% ^40,41^. The study sites were fixed, and outreach clinics in these two districts were run by the Liver Foundation, West Bengal. Patients presenting with a chief complaint of fever, shortness of breath, or musculoskeletal pain, or a known diagnosis of hypertension or diabetes mellitus were recruited for the study.

#### Participants

Eight nurses were hired using the following criteria: (a) completion of a B.Sc. in Nursing or General Nursing and Midwifery course, (b) proficiency in Bengali, (c) familiarity with smartphone use, and (d) ability to list two drugs for hypertension and for diabetes mellitus and two history questions for fever and for breathlessness.

#### Training

All nurses received forty hours of training over four weeks. A team of three physicians conducted training on history taking, medical conditions relevant to the study, diagnostic reasoning, the basics of artificial intelligence, and interacting with general-purpose LLM chats (ChatGPT and Gemini). Between didactic sessions, nurses practiced clinical and AI skills through patient role-play.

### Design approach

We followed the key activities of the HCD process defined in ISO 9241-210: understanding the context of use, specifying user requirements, producing design solutions, and evaluating the designs^42^, using established HCD research techniques^42,43^. To understand the context of use, two physicians performed a task analysis, decomposing a physician consultation into component tasks of history and examination, clinical reasoning, and patient communication, and mapping the consultation workflow as a process flow diagram. From this, we specified two user requirements: the solution had to be easy and acceptable for nurses to use during a consultation, and its clinical output had to be comparable in quality to a physician’s output. The design team developed successive prototypes, which nurses used in role-plays of simulated cases during training. Physician trainers observed these sessions as informal usability testing, and observations and nurse feedback informed each iteration. The penultimate and final versions reported in this paper were used in real patients.

### 4.4 Technical specifications

The NurseAI app is an Android application built with React Native and used on 5G smartphones running Android 14 or later. The phone recorded the interview and sent the audio over an encrypted (HTTPS) connection to a backend server on Amazon Web Services (AWS) Elastic Beanstalk. Audio recordings, Gemini-generated transcripts, and user authentication data were stored in a Supabase database. The backend attached fixed prompts and called a Gemini model through the Gemini API without an intermediate speech-to-text step; the model returned an English transcript of the Bengali audio with each response. We logged latency (wall-clock time from request to complete response), token usage, and cost for each call. The prompts and model specifications are available in our Zenodo repository.

### 4.5 Data privacy

Data from real patient encounters were encrypted in transit and at rest on AWS and Supabase servers located in India. Gemini API was used under Google’s paid-service terms which exclude prompts and responses from model improvement^44^. The patient interview transcripts were manually scrubbed of identifying information before being sent to graders for evaluation.

### 4.6. Code Availability

To enable replicability, analysis code is available through our Zenodo repository. The code for the NurseAI app is not publicly available. The app’s architecture and workflow are described in Sections 4.4 and 2.2. We have shared the prompts and model specifications in our Zenodo repository, but the reproduction of outputs is subject to stochasticity and continued model availability.

### 4.7 Evaluation methodology

We developed two versions of the app during the HCD process and evaluated the technical and clinical performance of version 2, which was deployed in the randomized crossover trial. Technical metrics include latency per model call, token use per consultation, and total cost of model calls per consultation. For clinical evaluation we operationalized the FAVES (Fairness, Appropriateness, Validity, Effectiveness, Safety) framework to a binary, multi-axis score^45^. Appropriateness includes consideration of the patient’s socioeconomic and geographic context in its plan. Validity is the logical consistency of the app’s differential diagnosis and suggested treatment with the evidence collected from the patient interview. Safety focuses on the likelihood of severe physical harm if the app’s treatment suggestions were applied exclusively and fully.

The evaluation set consisted of 50 patient consultations recruited for the study. The interview transcript was machine-translated to English and verified by a native speaker. The transcript and the app-generated assessment and plan were provided to the graders. Two graders independently scored the app’s assessment and plan for appropriateness, validity, and safety and noted reasons if these were marked ‘no’. Both graders were physicians with postgraduate training in medicine and practiced in rural India for more than five years. Graders were blinded to each other’s initial assessments and met to reach a consensus grade for each item.

#### 4.6.1 Statistical tests

We report proportions of consultations rated appropriate, valid, and safe with 95% confidence intervals (CIs) calculated using the Clopper-Pearson exact method. Technical metrics are reported as means. Inter-rater agreement is reported as the proportion of cases in which both graders gave identical ratings on all three criteria before consensus. Analyses were conducted in R version 4.6.0.

## Supporting information

Supplementary File 1: DECIDE-AI checklist

## Data Availability

Patient audio, assessments, and plans are not available because of privacy concerns and consent restrictions. De-identified transcripts and grading data are available subject to approval by the local ethics review board and the clinical partner (IILDS). Case-level consensus grading data and per-case technical metrics are available through our Zenodo repository

https://doi.org/10.5281/zenodo.24268547

## 5. Data Availability

Patient audio, assessments, and plans are not available because of privacy concerns and consent restrictions. De-identified transcripts and grading data are available subject to approval by the local ethics review board and the clinical partner (IILDS). Case-level consensus grading data and per-case technical metrics are available through our <u>Zenodo repository</u>

## 6. Supplementary information

Supplementary Information: Completed DECIDE-AI reporting checklist.

## 9. Author contributions

P.J., R.K., An.A., S.N., A.C. and P.M. conceptualised the study, and P.J., An.A. and S.N. developed the methodology. P.J., P.B., Aa.A., A.M., A.J. and S.S. developed the software, and P.J., P.B. and S.S. curated the data. P.J., C.M., A.K.T. and An.A. conducted the investigation. T.B. and S.N. performed the formal analysis; T.B. prepared the visualisations; and T.B., An.A. and S.N. validated the results. P.J., An.A. and P.M. provided resources. P.J., An.A. and S.N. administered the project, and P.J., An.A., S.N., A.C. and P.M. supervised the work. An.A. acquired funding. P.J., T.B. and An.A. wrote the original draft, and P.J., T.B., C.M., R.K., An.A., S.N., A.C. and P.M. reviewed and edited the manuscript. All authors reviewed the manuscript and approved the final version.

